# Acquired hypothyroidism, iodine status and hearing impairment in adults: a pilot study

**DOI:** 10.1101/2024.06.06.24308572

**Authors:** Tereza Grimmichova, Ludmila Verespejova, Zuzana Urbaniova, Martin Chovanec, Martin Hill, Radovan Bilek

## Abstract

**Objectives:** Hearing impairment can have major impacts on behavior, educational attainment, social status, and quality of life. In congenital hypothyroidism, the incidence of hearing impairment reaches 35-50%, while in acquired hypothyroidism there is a reported incidence of 25%. Despite this, knowledge of the pathogenesis, incidence and severity of hearing impairment remains greatly lacking. The aim of our study was to evaluate hearing in patients with acquired hypothyroidism.

**Methods:** 30 patients with untreated and newly diagnosed peripheral hypothyroidism (H) and a control group of 30 healthy probands (C) were enrolled in the study. Biochemical markers were measured, including median iodine urine concentrations (IUC) ⍰g/L. The hearing examination included a subjective complaint assessment, otomicroscopy, tympanometry, transitory otoacoustic emission (TOAE), tone audiometry, and brainstem auditory evoked potential (BERA) examinations. The Mann-Whitney U test, Fisher’s Exact test and multivariate regression were used for statistical analysis.

**Results:** The H and C groups had significantly different thyroid hormone levels (medians with 95% CI) TSH mU/L 13.3 (8.1, 19.3) vs. 1.97 (1.21, 2.25) p=0 and fT4 pmol/L 10.4 (9.51, 11.1) vs. 15 (13.8, 16.7) p=0. The groups did not significantly differ in age 39 (34, 43) vs. 41 (36,44) p=0.767 and IUC 142 (113, 159) vs. 123 (101, 157) p=0.814. None of the hearing examinations showed differences between the H and C groups: otomicroscopy (p=1), tympanometry (p=1), TOAE (p=1), audiometry (p=0.179), and BERA (p=0.505).

**Conclusions:** We did not observe any hearing impairment in adults with acquired hypothyroidism, and there were no associations found between hearing impairment and the severity of hypothyroidism or iodine status. However, some forms of hearing impairment, mostly mild, were very common in both studied groups.

## Introduction

Hypothyroidism can occur in countries that have sufficient iodine supplementation, with autoimmune thyroiditis the most common cause [1]. Patients with hypothyroidism report hearing losses; however, hearing impairments are poorly recognized and underreported, and often explicitly associated with congenital hypothyroidism and iodine deficiency [2]. It is well known that hearing deficits have a great impact on the education skills and social status of children and adolescents [3]. Youth with hearing impairments experience behavioral problems and demonstrate lower performance in oral language compared with peers with normal hearing [4]. Unfortunately, only limited studies have been dedicated to acquired hypothyroidism and hearing disabilities in adults [2, 5, 6].

The development of ears and their functioning depends on thyroid hormones. Thyroid hormones are necessary for maturation of the cochlea and the central auditory areas [7]. In humans, the critical period for hearing maturation corresponds approximately to an interval ranging from the early embryonic period to the first year of postnatal life [2]. Hypothyroidism occurring during this critical time window can lead to irreversible hearing impairment [8, 9]. However, when thyroxine (T4) therapy is begun early enough in life, it might still successfully reverse hearing loss [10]. Iodine is an essential component of the thyroid hormones triiodothyronine (T3) and thyroxine (T4). Thyroid hormone receptors (TR), occurring as the two isoforms TRα and TRβ, regulating thyroid hormone metabolism at the transcriptional level are expressed in the cochlea [11, 12]. Genetic disorders such as Pendred syndrome [13], thyroid hormone resistance [14], and thyroid hormones monocarboxylate transporter 8 abnormalities [15] support the links between thyroid hormones and the auditory system. The most well-known is Pendred syndrome, caused by biallelic mutations in the *SLC26A4/PDS* gene, which encodes the multifunctional anion exchanger pendrin. Pendred syndrome is an autosomal recessive disorder that is classically defined by the combination of sensorineural deafness/hearing impairment, goiter, and an abnormal organification of iodide with or without hypothyroidism [13, 16]. Importantly, Pendred syndrome accounts for about 4–10% of all cases of hereditary deafness. The hallmark of the syndrome is impaired hearing, which is associated with inner ear malformations such as an enlarged vestibular aqueduct [17, 18]. The organification defect is only mild and only seems to result in hypothyroidism under iodine-deficient conditions. Therefore, it remains unclear whether the sensorineural hearing loss associated with Pendred syndrome is solely caused by hypothyroidism [12, 13, 18].

Overall, it seems that hearing impairment associated with acquired hypothyroidism are most often bilateral, symmetrical, and sensorineural (perceptive), with some studies also supporting a conductive loss, and reversible to varying degrees after levothyroxine replacement therapy [6, 19]. Bircher was the first to describe the association of hearing impairment in patients suffering from a goiter as early as 1883 [20]. Various investigators have reported an association between thyroid hormone concentrations and hearing function in humans, but many of these studies are from the last century. In the 1950s, a study by Howarth and Lloyd 1956 described deafness under hypothyroidism as perceptive in type, with hypertrophy and edema of the mucosa of the nose, middle ear and eustachian tubes causing eustachian obstruction and thickening of the tympanic membrane. The study included seven females aged from 31-75 years with various levels of conductive hearing impairments (slight 0-20 dB loss to very severe perceptive deafness over 60 dB loss) and demonstrated improvements in some patients after thyroid hormone substitution [21]. In the 1970s the study Bhatia et al. 1977 confirmed hypothyroidism using estimates of serum protein-bound iodine in 43% of patients (total 72 patients) who had mild hearing loss as assessed by pure tone audiometry [22]. In the 1980s, twenty hypothyroid patients were investigated, demonstrating hearing losses in 80% of patients when compared with randomly selected age- and sex-matched normal subjects. Following treatment with levothyroxine a statistically significant improvement in hearing thresholds was observed by pure-tone audiometry. The investigators suggested a causal relationship between hypothyroidism and hearing loss [5]. In 2002, Malik et al. also reported a higher degree of hearing impairments in forty-five hypothyroid patients (age range 10–57 years). Pure tone audiometry (PTA) revealed hearing impairments in thirty-two patients, out of which fifteen had conductive, nine had mixed and eight had sensorineural impairments. Hearing impairment was correlated with increasing serum TSH levels and decreasing serum T3 and T4 levels (p > 0.05). Brainstem Evoked Response Audiometry (BERA) was performed in patients having hearing impairments shown by PTA. The interpeak interval (I-V) was >4.0 ms in 81.80% of ears, and the waves were not well formed with lower amplitude. After treatment with levothyroxine, hearing thresholds were significantly better in 30% of ears, with conductive impairment more likely to be improved [6]. This is in agreement with the findings of Bhatia et al 1977 who observed that the occurrence of hearing impairment gradually increased with an increase in the severity of hypothyroidism. The vestibular system was found to be affected only minimally [22]. Similar results but to a lesser extent were shown in a study by Aggarwal et al., with only about 13% cases having an objective audiological improvement, though many cases claimed a subjective improvement in hearing after thyroid hormone substitution. One case also had a deterioration in hearing [23]. In contrast, some studies have shown no hearing impairment in hypothyroid patients [24, 25] and others have not supported associations between hearing impairment and thyroid hormone levels in hypothyroid patients [26, 27].

Further, the evidence does suggest that iodine deficiency is related to hearing loss, and that supplementation in iodine-deficient individuals may improve hearing thresholds. Auditory impairment due to hypothyroidism causing such an iodine deficiency might exist [3]. The National Health and Nutrition Examination Survey is a cross-sectional, nationally representative survey of the noninstitutionalized civilian population of the United States analyzing data from 1198 adolescent (aged 12-19 years) participants. Urinary iodine concentration (UIC) less than 100 μg/L was found to be a predictive risk factor for having speech-frequency hearing loss among adolescents and more specifically among those with UICs less than 50 μg/L. However, the geometric mean of thyrotropin concentration of 1.4 *μ*UI/mL was in the normal range [28, 29]. In adults however, even less is about acquired hypothyroidism in relation to auditory function.

The aims of our study were to assess:

1. hearing impairment in relation to acquired hypothyroidism and associated changes to thyroid hormone levels.
2. hearing impairment due to iodine status as measured by median urinary iodine concentrations.
3. the risks of hearing loss and associations with the severity of hypothyroidism, including whether or not the severity of deafness is proportional to the degree of hypothyroidism and/or urinary iodine concentrations.

## Patients and Methods

The protocol of this prospective study complied with the Declaration of Helsinki and was approved by the Ethic Committee of the Faculty Hospital Kralovske Vinohrady. Before entering the study, written informed consent was obtained by the investigator from patients after they received both written and oral information. Patients’ history, medical records, ultrasound of the neck (US), and biochemical testing were done at two workplaces: the Institute of Endocrinology and the Department of Internal Medicine of the University Hospital Kralovske Vinohrady in Prague. Hearing tests were done at the Department of Otorhinolaryngology of the University Hospital Kralovske Vinohrady in Prague. A total of 30 hypothyroid patients and 30 matched healthy probands without history/treatment of thyroid disease were enrolled in the study [30]. The patients were all from the Czech Republic, a country with iodine sufficiency, and were consecutively recruited from 25^th^ May 2021 to 21^st^ February 2024. The inclusion and exclusion criteria are given in the study flow chart in Figure 1.

**Figure 1.**
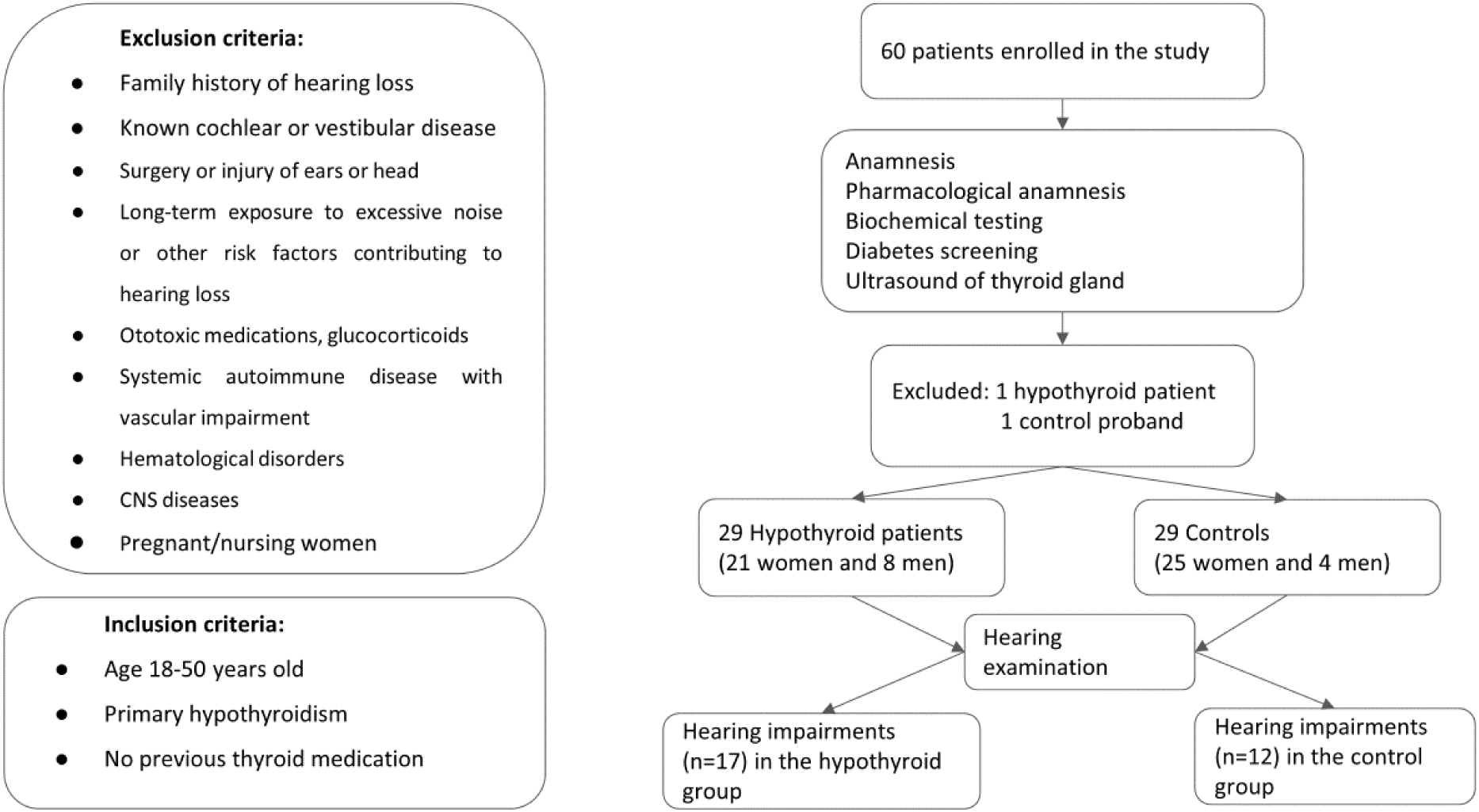
Flow chart of the study.

Overt primary hypothyroidism is defined as thyroid-stimulating hormone (TSH) concentrations above the reference range and free thyroxine (T4) concentrations below the reference range. The diagnosis of overt hypothyroidism was confirmed by a second blood sample. All the probands underwent the same study protocol with the same examinations. Biochemical testing consisted of measurement of electrolytes, liver and kidney tests, blood count, lipids, glucose, and Hb1ac to exclude diabetes, dyslipidemia, and systemic disease with impact on hear loss. Three consecutive morning urine samples were taken to measure median urinary iodine concentrations (UICs). UIC is an indicator of iodine status and reflects the current dietary intake of iodine. Urinary iodine below 20 μg/L denotes severe iodine deficiency, between 20–49 moderate, between 50–99 mild iodine deficiency, UIC between 100–199 is adequate iodine intake, UIC between 200–299 more than adequate, and UIC more than 300 μg/L is excessive iodine intake [31]. The WHOQOL-BREF questionnaire was used to evaluate the quality of life [32]. The hearing examination consisted of descriptions of subjective symptoms (tinnitus, dizziness, feeling of hear loss, and balance problem) and objective hearing exams: tympanometry (Maico MI 24), pure tone audiometry (Clinical Audiometer AC 40 Interacoustic), transitory otoacoustic emission (TOAE) (Titan Interacoustic) and Brain Evoked Response Audiometry (BERA) (Eclipse Interacoustic). Hearing losses were classified as either: mild (20 - 34 dB); moderate (35 - 49 dB); moderately severe (50 - 64 dB), severe (65-79 dB), profound (80-94 dB), or as complete hearing loss (> 95 dB). The Global Burden of Disease (GBD) scale defines hearing loss as the quietest sound an individual can hear in their better ear, taken as the pure-tone average (PTA) of audiometric thresholds of 0.5 kHz, 1 kHz, 2 kHz, and 4 kHz [33, 34]. Serum TSH (0.270-4.200 mIU/L), fT4 (11.9-21.6 pmol/L), fT3 (3.10-6.80 pmol/L), TRAbs (0.00-1.75 IU/L), thyroglobulin (3.50-77.00 ug/L) concentrations were measured using the ECLIA method (Roche). The HbA1C test was performed using an ion exchange HPLC method that is certified by the NGSP (www.njsp.org) and standardized or traceable to the Diabetes Control and Complications Trial (DCCT) reference assay. Serum anti-Tg (0.01-120.00 IU/mL) and anti-TPO (0.01-40.00 IU/mL) were measured by ELISA (Aeskulisa). Urinary iodine concentrations were measured by absorption spectrophotometry at a wavelength of 430 nm. Glucose was measured by a spectrophotometry (UV)-hexokinase method. Liver enzymes and lipids were measured by absorption spectrophotometry (Cobas PRO, Roche), and blood count (Celtac F, Nihon Kohden).

## Statistics

Statistical significance was defined as p-values <0.05. The Mann-Whitney U test ((medians with 95% CI) and Fisher’s Exact test were used for statistical analysis. The importance of individual predictors in discriminating between individual groups was evaluated using multivariate regression with a reduction of dimensionality known as orthogonal projections to latent structure (OPLS) for one predicted (dependent) variable in the model [35].

## Results

A total of 60 patients were enrolled in the study, but two patients had to be excluded: one hypothyroid patient due to newly diagnosed severe iron deficiency anemia and one control proband due to newly diagnosed polycythemia vera. Finally, 29 hypothyroid patients (H; 21 women and 8 men) and 29 controls (C; 25 women and 4 men) were statistically analyzed. The H and C groups were not statistically different in either age (medians with 95% CI) 39 (34, 43) vs. 41 (36, 44) (p=0.767) or gender (p=0.331). The H group had significantly higher levels than the C group of: TSH 13.3 (8.1, 19.3) vs. 1.97 (1.21, 2.25) mU/L (p=0); anti-TPO 335 (164, 520) vs. 28 (4.78, 31) IU/mL (p < 0.01); and anti-Tgl IU/mL 32.6 (4.88, 58.6) vs. 1.3 (1.3, 1.3) IU/mL (p<0.001). The H group had significantly lower fT4 10.4 (9.51, 11.1) vs. 15 (13.8, 16.7) pmol/L (p=0) and fT3 4.61 (4.35, 5.1) vs. 5.24 (5.07, 5.5) pmol/L (p=0.009) compared to the C group. For more details see Table 1.

**Table 1.**
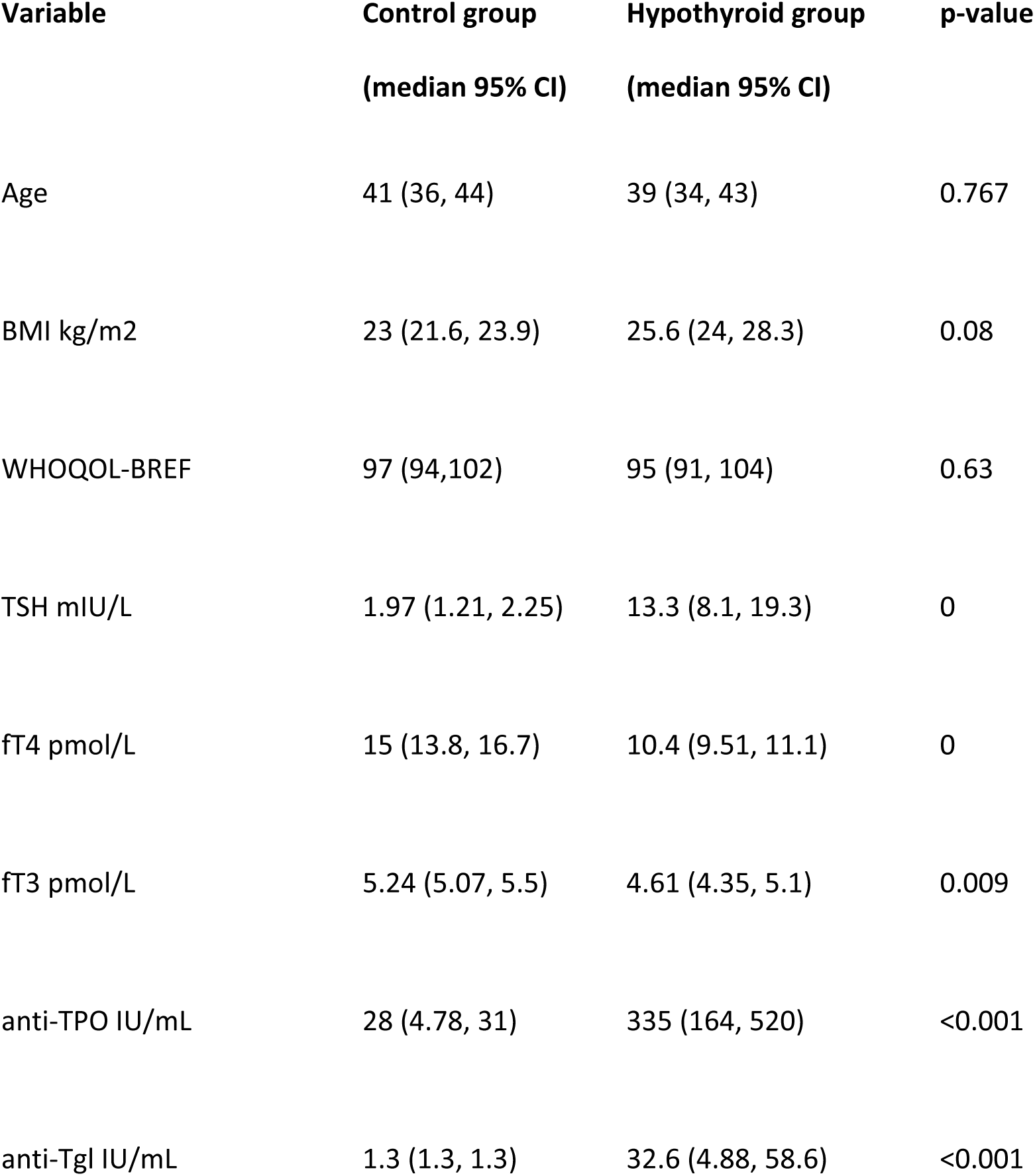
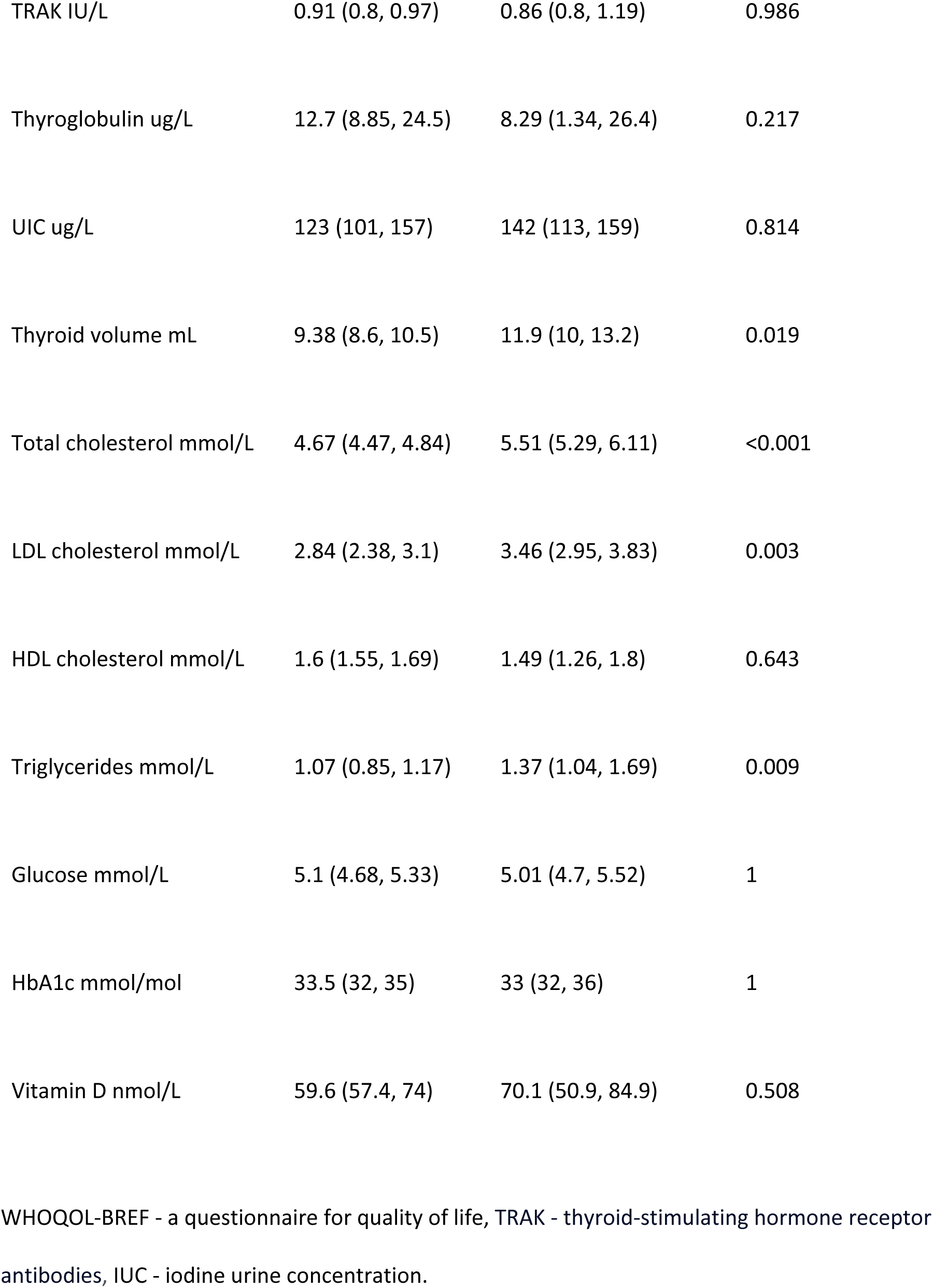
The Mann-Whitney test was used to compare the control and hypothyroid groups.

The cause of hypothyroidism was autoimmune thyroiditis (p=0). The H and C groups were comparable in IUCs 142 (113, 159) vs. 123 (101, 157) ⍰g/L (p=0.814). Only 15 probands had mild iodine deficiencies, with the lowest IUC of 50 ⍰g/L. For more details see Table 2.

**Table 2.** Fisher’s exact 2-sided test was used to compare the control and hypothyroid groups.

| Variable | Control group | Hypothyroid group | p-value |
| --- | --- | --- | --- |
| Male | 13.8% | 27.6% | 0.331 |
| Smoking | 3.7% | 18.2% | 0.160 |
| Autoimmune thyroiditis | 3.4% | 86.2% | 0 |
| Otomicroscopy | 0% | 3.4% | 1 |
| Tympanometry | 6.9% | 6.9% | 1 |
| Audiometry | 27.6% | 10.3% | 0.179 |
| GBD R | 3.4% | 0% | 1 |
| GBD L | 3.4% | 0% | 1 |
| BERA | 13.8% | 21.4% | 0.505 |

GBD R - GBD scale right ear, GBD L - GBD scale left ear, BERA - brainstem evoked response audiometry.

Out of the 58 patients in the study group, only 3 patients in the C group presented with subjective complaints, including unilateral hearing loss and tinnitus. There were no subjective complaints in the H group (p=0.239). Otomicroscopy revealed no pathologies except for one case of an atrophic eardrum (p=1). Some hearing impairment in the patients was observed in 29 patients, of which 17 were in the hypothyroid group and 12 control probands. Hearing impairments predominantly affected high-tone frequencies (6 kHz and 8 kHZ) in the range from 20-45dB. We did not observe any deficiency of more than 45 dB. The hearing examinations tympanometry (p=1), TOAE (p=1), audiometry (p=0.179), and BERA (p=0.505) were not different between the H and C groups. For more details see Table 2 and 3.

**Table 3.** The Mann-Whitney test was used to compare the control and hypothyroid groups.

| Variable | Control group<br>(median 95% CI) | Hypothyroid group<br>(median 95% CI) | p-value |
| --- | --- | --- | --- |
| PTA R (dB) | 9 (8,10) | 8 (6,9) | 0.120 |
| PTA L (dB) | 9 (8, 10) | 9 (6,10) | 0.456 |
| Audiometry 6 kHz R (dB) | 10 (10, 15) | 10 (10,15) | 0.833 |
| Audiometry 6 kHz L (dB) | 10 (10, 20) | 15 (10,15) | 0.768 |
| Audiometry 8kHz R (dB) | 10 (10,15) | 15 (10, 20) | 0.387 |
| Audiometry 8kHz L (dB) | 10 (10, 20) | 15 (10,20) | 0.880 |

PTA R - pure-tone average right ear, PTA L - pure-tone average left ear. R - right ear, L - left ear.

The OPLS statistical analysis used to evaluate the importance of individual predictors to discriminate between the hypothyroid and control groups found no statistically significant predictors of hearing impairments.

## Discussion

In our study, we did not observe any hearing impairments related to acquired hypothyroidism in adults. Further our data did not support any associations between hearing impairment and the degree of hypothyroidism and iodine status. However, some forms of hearing impairment, mostly mild, were very common in both studied groups.

The prevalence of overt hypothyroidism in the general population varies between 0–2% and 3-5% in Europe, and the prevalence of undiagnosed hypothyroidism, including both overt and mild cases, is around 5%. In iodine-sufficient areas, the most common cause of hypothyroidism is chronic autoimmune thyroiditis [36]. We concur with these results and we have to point out that we had some difficulties finding a patient with manifest hypothyroidism. Only three patients had TSH levels over 50 mU/L. The probable reason was that our patient cohorts were younger, but widespread thyroid function testing and relatively low thresholds for treatment initiation almost certainly also play a role [37].

Hearing impairment is a major public health problem and is one of the leading causes of years lived with a disability. Moreover, according to the World Health Organization, the incidence of auditory disorders is increasing at an alarming rate without much concern or perception by society or public health officials [38]. Hearing impairments can affect the quality of life from childhood, with poor school performance common, as well as in adulthood, with higher risks of unemployment or lower earnings [39]. Hearing impairment is also associated with increased cognitive dysfunction and dementia in the elderly [40]. There are many reasons for hearing impairment in adults and it is strongly associated with age. We chose to study younger probands (<50 years old) in order to eliminate the risks of presbycusis, metabolic disorders such as type 2 diabetes, obesity, hypertension and dyslipidemia with their associated negative effects on hearing [39, 41]. Not surprisingly, our hypothyroid group of probands had higher lipids levels compared to the control group, but without any impact on hearing impairment. The majority of probands were non-smokers, so this risk factor was not relevant in this study. In comparison to some other studies [6, 22, 5] we can just speculate on the divergent results. Most of the studies were done in the past, and recent studies are mostly from India and the Middle East [26, 42, 43]. This geographical variation could introduce several confounding factors, including the prevalence of occupational noise exposure, preventable infections such as chronic otitis media and meningitis, nutritional status and health-care access. Further, there is evidence of improvements in hearing between 1959–1962 and 1999–2004, suggesting a beneficial trend for at least half a century. Explanations for this trend could include reductions in occupational noise exposure, less smoking, and better management of other cardiovascular risk factors such as hypertension and diabetes [33].

Previous studies were also frequently done with a small number of probands with ages ranging from 12 to 75 years old. Further, the hypothyroid status of probands tended to be very heterogeneous, from subclinical forms to severe myxedema [44]. In the study of Malik et al., 2002, moderate hearing impairment was present approximately in 48% and mild impairment in 41% of ears, whereas severe or profound hearing impairment was not found in any case [6]. In agreement with previous studies, we observed mostly mild and only two cases with moderate hearing impairments in both hypothyroid and control groups. In later studies, hearing examinations were usually more detailed than thyroid testing and evaluations of iodine status [6, 42, 26]. Some other nutritional factors such as single nutrients (vitamin A, B, C, D and E, and zinc, Mg, Se, iodine, and iron) are associated with hearing status. Serum thyroglobulin is well correlated with the severity of iodine deficiency as measured by median urinary iodine concentrations (UICs) [45], but our groups had comparable levels of both thyroglobulin and UICs in the optimal range. Based on WHO data, the number of countries with iodine insufficiency has declined from 113 (estimated by total goiter rates) in 1993 to 20 (estimated by urinary iodine concentrations) in 2017 [46]. Congenital iodine deficiency can be manifested as two syndromes: a more common neurological disorder with brain damage, deaf mutism, squint and spastic paresis of the legs. Such patients are usually euthyroid, but goiter and hypothyroidism can be seen in some cases. Urinary iodine levels are usually less than 20 ⍰g/L. The less common syndrome includes severe hypothyroidism and growth retardation but a less severe mental defect. Both conditions are due to dietary iodine deficiency and can be prevented by iodine supplementation before pregnancy [47]. Hearing loss has also been observed in adolescents (aged 12-19 years) associated with iodine deficiency in spite of normal TSH levels [28]. Therefore, it remains uncertain whether the hearing loss is solely caused by hypothyroidism or iodine deficiency. It appears that iodine plays a role in immune responses and might have a beneficial influence on mammary dysplasia and fibrocystic breast disease [48]. It is possible that iodine itself could have some other roles in human physiology.

One of the limits of our study is the low number of probands, but this was rather a pilot study about a topic with very limited evidence. We also did not retest the patients after thyroxine replacement therapy reaching euthyroidism, since there were no significant differences between the hypothyroid and control groups in hearing impairments. There are some previous studies showing significant improvement of hearing threshold in 12.5 up to 73% ears at several levels, middle ear, cochlear or retro-cochlear [42]. Bhatia et al 1979 reported definite subjective improvement in hearing on becoming euthyroid, but this was not confirmed by audiometry. Another limit of our study could be that the hypothyroid state was not too advanced in our patients. All of our hypothyroid patients complied with the definition of hypothyroidism, i.e. increased levels of TSH and simultaneously decreased fT4 levels in spite of fT3 being in normal range in some of the probands. There are adaptive mechanisms during iodine deficiency, or the incipient phase of hypothyroidism, preserving optimal plasma and tissue T3 levels linked to deiodinase 2 expression. The signs and symptoms of overt hypothyroidism are minimized by preserving T3 levels. Thus, serum T3 levels perform poorly as a tool to diagnose hypothyroidism compared to serum FT4 or TSH. Nevertheless, it is clear that plasma and tissue T3 contents are associated and that changes in plasma T3 are directly connected to changes in the tissue T3 content [49]. We cannot exclude that patients with parallel drops in T4 and T3 levels could be at risk of hearing impairments, but our statistical analysis did not observe any links between thyroid hormone levels and such impairment. A benefit of our study is the prospective design with comprehensive examinations of patients and excluding more common causes of hearing loss. We included evaluations of iodine status, which has a crucial effect on hearing impairment. Finally, the quality of life of our probands established by the WHOQOL-BREF questionnaire was comparable for both groups.

## Conclusions

We did not observe any hearing impairment in adults associated with acquired hypothyroidism. However, some forms of hearing impairment, mostly mild, were very common in both studied groups. Since our study did not show any associations between hearing impairment and either the severity of hypothyroidism or iodine status, we suppose that there are more important causes of hearing impairment than hypothyroidism in adults.

## Data Availability

All relevant data are within the manuscript.

## Acknowledgements

The authors acknowledge D. Hardekopf for proofreading activity.

## References

1. Ragusa F, Fallahi P, Elia G, Gonnella D, Paparo SR, Giusti C, et al. Hashimotos’ thyroiditis: Epidemiology, pathogenesis, clinic and therapy. Best Pract Res Clin Endocrinol Metab. 2019;33: 101367.

2. Melse-Boonstra A, Mackenzie I. Iodine deficiency, thyroid function and hearing deficit: a review. Nutr Res Rev. 2013;26: 110–7.

3. Zimmermann MB. The adverse effects of mild-to-moderate iodine deficiency during pregnancy and childhood: a review. Thyroid. 2007;17: 829–835.

4. Clark C, Sörqvist P. A 3 year update on the influence of noise on performance and behavior. Noise Health. 2012;14: 292–296.

5. Anand VT, Mann SB, Dash RJ, Mehra YN. Auditory investigations in hypothyroidism. Acta Otolaryngol. 1989;108: 83–7.

6. Malik V, Shukla GH, Bhatia N. Hearing profile in hypothyroidism. Indian J Otolaryngol Head Neck Surg. 2002; 54: 285–290.

7. Sohmer H, Freeman S. The importance of thyroid hormone for auditory development in the fetus and neonate. Audiol Neurootol. 1996;1: 137–147.

8. Graven SN, Browne JV. Auditory development in the fetus and infant. Newborn Infant Nurs Rev. 2008;8: 187–193.

9. Knipper M, Richardson G, Mack A, Müller M, Goodyear R, Limberger A, et al. Thyroid hormone-deficient period prior to the onset of hearing is associated with reduced levels of β-tectorin protein in the tectorial membrane. Implications for hearing loss. J Biol Chem. 2001; 276: 39046–39052.

10. Wasniewska M, De Luca F, Siclari S, Salzano G, Messina MF, Lombardo F, et al. Hearing loss in congenital hypothalamic hypothyroidism: a wide therapeutic window. Hear Res. 2002;172: 87–91.

11. Bradley DJ, Towle HC, Young WS. Alpha and beta thyroid hormone receptor (TR) gene expression during auditory neurogenesis: evidence for TR isoform-specific transcriptional regulation in vivo. Proc Natl Acad Sci U S A. 1994;91: 439–443.

12. Forrest, D. Deafness and goiter: molecular genetic considerations. J Clin Endocrinol Metab. 1996; 81: 2764–2767.

13. Bizhanova A, Kopp P. Controversies concerning the role of pendrin as an apical iodide transporter in thyroid follicular cells. Cell Physiol Biochem. 2011;28: 485–490.

14. Brucker-Davis F, Skarulis MC, Grace MB, Benichou J, Hauser P, Wiggs E, et al. Genetic and clinical features of 42 kindreds with resistance to thyroid hormone. The National Institutes of Health Prospective Study. Ann Intern Med. 1995;123: 572–583.

15. Dumitrescu AM, Liao XH, Best TB, Brockmann K, Refetoff S. A novel syndrome combining thyroid and neurological abnormalities is associated with mutations in a monocarboxylate transporter gene. Am J Hum Genet. 2004;74: 168–75.

16. Coakley JC, Keir EH, Connelly JF. The association of thyroid dyshormonogenesis and deafness (Pendred syndrome): experience of the Victorian Neonatal Thyroid Screening Programme. J Paediatr Child Health. 1992;28: 398–401.

17. Bogazzi F, Russo D, Raggi F, Ultimieri F, Berrettini S, Forli F, et al. Mutations in the SLC26A4 (pendrin) gene in patients with sensorineural hearing loss and enlarged vestibular aqueduct. J Endocrinol Invest. 2004;27: 430–435.

18. Wémeau JL, Kopp P. Pendred syndrome. Best Pract Res Clin Endocrinol Metab. 2017;31: 213–224.

19. Comer DM, McConnell EM. Hypothyroid-associated sensorineuronal deafness. Ir J Med Sci. 2010;179: 621–2.

20. Bircher H. Der Endemische Kropf. Basel 1883.

21. Howarth AE, Lloyd HE. Perceptive deafness in hypothyroidism. Br Med J. 1956;1: 431–3.

22. Bhatia L, Gupta OP, Agrawal MK, Mishr SK. Audiological and vestibular function tests in hypothyroidism. Laryngoscope 1977;87: 2082–9.

23. Aggarwal MK, Singh GB, Nag RK, Singh SK, Kumar R, Yadav M. Audiological Evaluation in Goitrous Hypothyroidism. Int J Otolaryngol Head Neck Surg. 2013;2: 201–206.

24. Post JT. Hypothyroid deafness. A clinical study of sensori-neural deafness associated with hypothyroidism. Laryngoscope. 1964;74: 221–32.

25. DeVos JA, Parving A, Ostri B, Bretlau P, Hansen JM, Parving HH. Audiological and temporal bone findings in myxedema. Ann Otol Rhinol Laryngol. 1986;95: 278–83.

26. Balaji PV, Thirumaran M, Sharathbabu V. Assessment of audiologic evaluation in patients with acquired hypothyroidism. IAIM 2016; 3(7): 222–227.

27. Santos KT, Dias NH, Mazeto GM, Carvalho LR, Lapate RL, Martins RH. Audiologic evaluation in patients with acquired hypothyroidism. Braz J Otorhinolaryngol. 2010;76: 478–84.

28. Scinicariello F, Buser MC. Association of Iodine Deficiency with Hearing Impairment in US Adolescents Aged 12 to 19 Years: Analysis of NHANES 2007-2010 Data. JAMA Otolaryngol Head Neck Surg. 2018;144: 644–645.

29. Millon-Ramirez C, García-Fuentes E, Soriguer F. Iodine Deficiency and Hearing Impairment. JAMA Otolaryngol Head Neck Surg. 2019;145:94–95.

30. Zamrazil V, Bilek R, Cerovska J, Delange F. The elimination of iodine deficiency in the Czech Republic: the steps toward success. Thyroid 2004;14: 49–56.

31. World Health Organization 2007. Assessment of Iodine Deficiency Disorders and Monitoring Their Elimination: A Guide for Programme Managers 3rd ed. Geneva, Switzerland: World Health Organization; 2007. Available from: http://apps.who.int/iris/bitstream/10665/43781/1/9789241595827_eng.pdf.

32. Skevington SM, Lotfy M, O’Connell KA; WHOQOL Group. The World Health Organization’s WHOQOL-BREF quality of life assessment: psychometric properties and results of the international field trial. A report from the WHOQOL group. Qual Life Res. 2004;13: 299–310.

33. Hoffman HJ, Dobie RA, Losonczy KG, Themann CL, Flamme GA. Declining prevalence of hearing loss in US adults aged 20 to 69 years. JAMA Otolaryngol Head Neck Surg. 2017;143: 274–285.

34. GBD 2019 Hearing Loss Collaborators. Hearing loss prevalence and years lived with disability, 1990-2019: findings from the Global Burden of Disease Study 2019. Lancet 2021;397: 996–1009.

35. Meloun M, Hill M, Militky J, Kupka K. Transformation in the PC-aided biochemical data analysis. Clin Chem Lab Med. 2000;38: 553–9.

36. Chaker L, Bianco AC, Jonklaas J, Peeters RP. Hypothyroidism. Lancet. 2017 23;390: 1550–1562.

37. Taylor, P., Albrecht, D., Scholz, A. et al. Global epidemiology of hyperthyroidism and hypothyroidism. Nat Rev Endocrinol. 2018;14: 301–316.

38. Graydon K, Waterworth C, Miller H, Gunasekera H. Global burden of hearing impairment and ear disease. J Laryngol Otol. 2019;133: 18–25.

39. Jung SY, Kim SH, Yeo SG. Association of Nutritional Factors with Hearing Loss. Nutrients. 2019;11: 307.

40. Lin FR, Yaffe K, Xia J, Xue QL, Harris TB, Purchase-Helzner E, et al. Hearing loss and cognitive decline in older adults. JAMA Intern. Med. 2013;173: 293–299.

41. Kim SH, Won YS, Kim MG, Baek YJ, Oh IH, Yeo SG. Relationship between obesity and hearing loss. Acta Otolaryngol. 2016;136: 1046–1050.

42. Singh R, Aftab M, Jain S, Kumar D. Audiological Evaluation in Hypothyroid Patients and Effect of Thyroxine Replacement Therapy. Indian J Otolaryngol Head Neck Surg. 2019;71: 548–552.

43. Mahfzah M, Alhawari H, Momani SM, Abbasi H. The prevalence of hearing loss in patients with autoimmune thyroid disease: A prospective study. Jordan Medical Journal. 2018; 52: 109–116.

44. Vant Hoff W, Stuard DW. Deafness in myxedema. O J Med. 1979; 48: 361–7.

45. Knudsen N, Bulow I, Jørgensen T, Perrild H, Ovesen L, Laurberg P. Serum Tg: a sensitive marker of thyroid abnormalities and iodine deficiency in epidemiological studies. J Clin Endocrinol Metab. 2001;86: 3599–3603.

46. Iodine Global Network (2016) Global scorecard 2016: moving toward optimal iodine status. Available from: http://www.ign.org/newsletter/idd_nov16_global_scorecard_2016.pdf.

47. Chen ZP, Hetzel BS. Cretinism revisited. Best Pract Res Clin Endocrinol Metab. 2010;24: 39–50.

48. Kessler JH. The effect of supraphysiologic levels of iodine on patients with cyclic mastalgia. Breast J. 2004;10: 328–336.

49. Salas-Lucia F, Bianco AC. T3 levels and thyroid hormone signaling. Front Endocrinol (Lausanne). 2022;13: 1044691.

